# Investigation of Vaccine Breakthrough Infections by Vaccine strategy during the Delta Variant Wave in France

**DOI:** 10.1101/2021.12.05.21267301

**Authors:** Antonin Bal, Grégory Destras, Bruno Simon, Jean-Marc Giannoli, Florence Morfin, Bruno Lina, Laurence Josset, IVAC study group

## Abstract

Herein, we describe the characteristics of vaccine breakthrough infections (VBI) in fully vaccinated individuals according to five vaccine strategies during the Delta wave in France. Inclusion criterion was a positive test at least 2 weeks after a full vaccine schedule: homologous vaccination with Pfizer-BioNTech (BNT162b2) or Moderna (mRNA-1273); heterologous vaccination with Astrazeneca and Pfizer-BioNTech (ChadOx1/BNT162b2); single-dose vaccines Johnson & Johnson (Ad26.COV2.S) or Astrazeneca (ChadOx1). A total of 1630 VBI from patients fully vaccinated between February and July were included in this study. SARS-CoV-2 sequencing performed for 1366 samples showed that the delta variant represented 94.1% (1286/1366). Delta-VBI were mainly symptomatic (mild symptoms) with no difference according to the vaccine strategy (p=0.362). The median RT-PCR Ct values at diagnosis were significantly different between symptomatic and asymptomatic cases only for BNT162b2 group (17.7 (15.07, 20.51) vs 19.00 (16.00, 23.00), p=0.004). Up to 50% of VBI was classified as early-VBI (infected less than one month after full immunization) for BNT162b2, mRNA-1273, ChadOx1, and J Ad26.COV2.S. People aged 14-49 yo were overrepresented in early VBI compared to non-early VBI for BNT162b2 and mRNA-1273 (73.92% vs 37.87% for BNT162b2 and 77.78% vs 46.67 % for mRNA-1273, p<0.05). Our data emphasize a high prevalence of Delta-VBI occurring only one month after full immunization in young patients that might be related to relaxation of barrier gestures.

---

As of 29 November 2021, more than 89% of the adult population was fully vaccinated with different vaccine strategies in France ^1^. During this campaign, Delta vaccine breakthrough infections (VBI) associated with a high viral load have been reported ^2^ Furthermore, level and duration of humoral and cellular immune responses was shown to be different according to the vaccine used ^3,4^. Herein, we describe the characteristics of VBI in fully vaccinated individuals according to five vaccine strategies during the Delta wave in France.

An observational study was conducted at the national reference center for respiratory viruses in Lyon, France from April 2021 to August 2021. During this period, samples positive for SARS-CoV-2 in Biogroup community testing laboratories were retrieved for whole genome sequencing. Inclusion criterion was a positive test at least 2 weeks after a full vaccine schedule: homologous vaccination with Pfizer-BioNTech (BNT162b2) or Moderna (mRNA-1273); heterologous vaccination with Astrazeneca and Pfizer-BioNTech (ChadOx1/BNT162b2); single-dose vaccines Johnson & Johnson (Ad26.COV2.S) or Astrazeneca (ChadOx1).

Ethics approval was obtained from the national review board for biomedical research (Comité de Protection des Personnes Centre-Ouest I, France; ID-RCB 2021-A01877-34), and the study was registered on ClinicalTrials.gov (NCT05060939). Continuous variables are presented as median with interquartile range (IQR) and compared using non-parametric Kruskal–Wallis tests. Proportions were compared using the chi-squared or Fisher’s exact test as appropriate. A p-value of <0.05 was regarded as statistically significant. Statistical analyses were conducted using R software, version 4.0.5 (R Foundation for Statistical Computing).

A total of 1630 VBI from patients fully vaccinated between February and July were included in this study. SARS-CoV-2 sequencing performed for 1366 samples showed that the delta variant represented 94.1% (1286/1366) of VBI (Figure 1A).

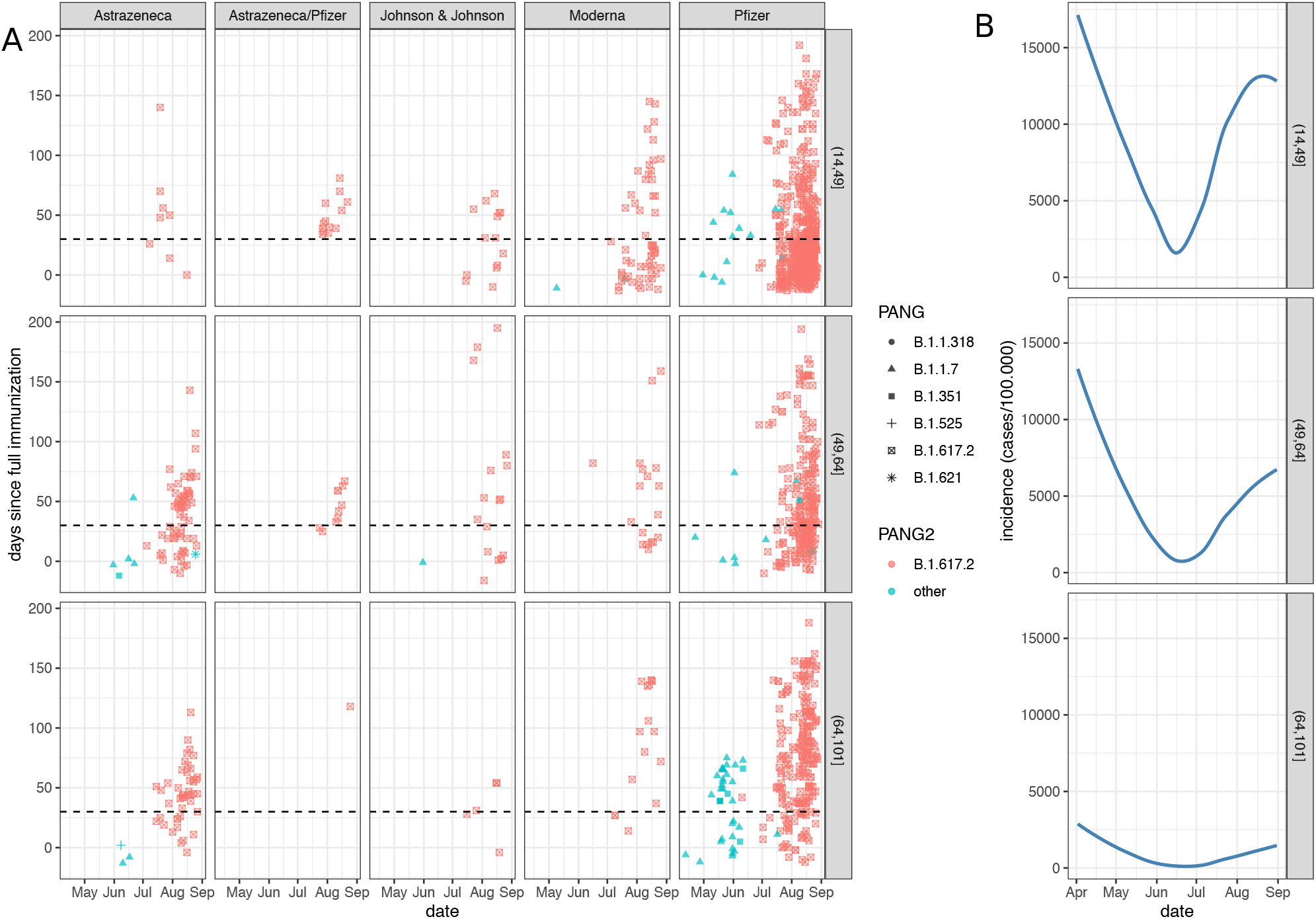
NGS-confirmed vaccine breakthrough infections in fully vaccinated individuals, April, 12^th^ to August, 30^th^. A. Vaccine breakthrough infections (VBI) are represented by vaccine strategy and by age group. The delay since full immunization are represented in the y-axis. The horizontal dotted lines indicate the delay of one month following the full immunization. VBI represented below these lines are defined as early VBI. Each Pango lineage determined by whole genome sequencing is represented by a symbol. Lineage B.1.617.2 (Delta variant) is represented in red. B. Incidence of COVID-19 cases in France is represented by age group ^6^.

Among delta-VBI, BNT162b2 was the most frequent vaccine reported (1017/ 79.1%) followed by ChadOx1 (118/1286, 9.2%) and mRNA-1273 (90/1286, 7.0%, Table). The number of VBI in each age group followed the dynamics of SARS-CoV-2 infections in France with a peak occurring in mid-August (Figure 1B). The median age of Delta-VBI cases significantly differed depending on the vaccine strategy with a median below 50 yo except for Ad26.COV2.S and ChadOx1 groups (p<0.001). Delta-VBI were mainly symptomatic (mild symptoms) with no difference according to the vaccine strategy (p=0.362). The median RT-PCR Ct values at diagnosis were significantly different between symptomatic and asymptomatic cases only for BNT162b2 group (17.7 (15.07, 20.51) vs 19.00 (16.00, 23.00), p=0.004).

Interestingly, more than 75% of Delta-VBI occurred in patients who were within three months following full immunization, regardless of the vaccination strategy. Besides, up to 50% of VBI was classified as early-VBI (infected less than one month after full immunization) for BNT162b2, mRNA-1273, ChadOx1, and J Ad26.COV2.S (Table). People aged 14-49 yo were overrepresented in early VBI compared to non-early VBI for BNT162b2 and mRNA-1273 (73.92% vs 37.87% for BNT162b2 and 77.78% vs 46.67 % for mRNA-1273, p<0.05, Figure 1).

Our data showed that mild VBI detected in community laboratories during the spread of Delta mainly occurred within three months after full immunization for all vaccine strategies. Low Ct values and presence of symptoms observed herein support a risk of transmission as previously noted in fully vaccinated health care workers infected by Delta variant ^5^.

The main limitations of this study are related to the French vaccine policies explaining the limited sample sizes for non-RNA vaccines and differences in vaccination date between age groups and vaccine types. Yet our data emphasize a high prevalence of Delta-VBI occurring only one month after full immunization in young patients that might be related to relaxation of barrier gestures. The surveillance of VBI should be reinforced in the context of the booster vaccination campaign.

**Table.** Characteristics of Delta vaccine breakthrough infections in fully vaccinated individuals by vaccine strategy.

|  | Astrazeneca<br>(ChadOx1)<br>N = 118 | Astrazeneca/<br>Pfizer<br>(ChadOx1/BNT162b2)<br>N = 24 | Johnson & Johnson<br>(Ad26.COV2.S)<br>N = 37 | Moderna<br>(mRNA-1273)<br>N = 90 | Pfizer-BioNTech<br>(BNT162b2)<br>N = 1017 | Total<br>N = 1,286 | p |
| --- | --- | --- | --- | --- | --- | --- | --- |
| **Symptomatic** |  |  |  |  |  |  |  |
| n (%) | 50/72<br>(69.44%) | 18/21 (85.71%) | 19/26 (73.08%) | 40/54 (74.07%) | 506/626<br>(80.83%) | 633/799<br>(79.22%) | 0.127 |
| **Ct at diagnosis** |  |  |  |  |  |  |  |
| symptomatic, median (IQR) | 17.30<br>(14.33, 20.00)<br>N=47 | 19.60<br>(15.13, 22.00)<br>N=17 | 16.55<br>(14.59, 21.90)<br>N=19 | 18.50 (15.34, 22.50)<br>N=37 | 17.74 (15.07, 20.51)<br>N= 358 | 17.74 (15.00, 21.00)<br>N=478 | 0.362 |
| asymptomatic, median (IQR) | 18.47<br>(16.00, 20.15)<br>N=22 | 18.30<br>(18.25, 19.65)<br>N=3 | 19.30<br>(17.05, 22.09)<br>N=7 | 21.88 (19.85, 23.12)<br>N=14 | 19.00 (16.00, 23.00)<br>N=90 | 19.55 (16.00, 23.00)<br>N=136 | 0.382 |
| p.value | 0.141 | 0.681 | 0.523 | 0.187 | 0.004 | 0 |  |
| **Comorbidities** |  |  |  |  |  |  |  |
| n (%) | 29/54 (53.70%) | 4/8 (50.00%) | 0/3 (0.00%) | 10/36 (27.78%) | 108/402<br>(26.87%) | 151/503<br>(30.02%) | 0.001 |
| **Sex** |  |  |  |  |  |  |  |
| Female n (%) | 65 (55.08%) | 16 (66.67%) | 18 (48.65%) | 47 (52.22%) | 590 (58.01%) | 736 (57.23%) | 0.496 |
| **Age** |  |  |  |  |  |  |  |
| median (IQR) | 62.00<br>(58.00, 68.00) | 45.00<br>(36.75, 53.00) | 57<br>(43.00, 62.00) | 41.00 (26.25, 57.00) | 46<br>(32.00, 61.00) | 49.00<br>(34.00, 62.75) | p<0.001 |
| **Time between full immunization and positive PCR** |  |  |  |  |  |  |  |
| All, median days (IQR) | 45.00<br>(21.25, 56.75) | 44.00<br>(36.00, 60.25) | 35.00<br>(6.00, 54.00) | 30.50<br>(10.00, 82.00) | 30.00<br>(11.00, 73.00) | 33.00<br>(12.00, 70.00) | 0.171 |
| 14-49yo, median days (IQR) | 63.00<br>(37.00, 73.50) | 57<br>(53.00, 74.00) | 45.00<br>(20.50, 66.00) | 34.00<br>(14.75, 80.25) | 33.00<br>(19.00, 54.00) | 34<br>(19.00, 59.00) | 0.007 |
| 50-64yo, median days (IQR) | 59.00<br>(33.00, 69.25) | 58.50<br>(47.75, 73.00) | 65<br>(19.00, 94.00) | 65.00<br>(31.00, 91.75) | 49.50<br>(32.75, 87.00) | 52<br>(33.00, 82.00) | 0.682 |
| >64, median days (IQR) | 46; 58.00<br>(41.00, 71.00) | 132<br>(132.00, 132.00) | 56.50<br>(42.75, 68.00) | 111.00<br>(66.00, 150.75) | 90.50<br>(56.00, 124.00) | 84<br>(53.00, 118.00) | 0 |
| **Cumulative breakthrough cases within** |  |  |  |  |  |  |  |
| 1 month n (%) | 43 (36.44%) | 2 (8.33%) | 15 (40.54%) | 45 (50.00%) | 510 (50.15%) | 615 (47.82%) | 0 |
| 2 months n (%) | 95 (80.51%) | 18 (75.00%) | 29 (78.38%) | 54 (60.00%) | 714 (70.21%) | 910 (70.76%) | 0.018 |
| 3 months n (%) | 113 (95.76%) | 23 (95.83%) | 34 (91.89%) | 71 (78.89%) | 846 (83.19%) | 1,087<br>(84.53%) | 0.001 |
| 4 months n (%) | 116 (98.31%) | 24 (100.00%) | 34 (91.89%) | 78 (86.67%) | 920 (90.46%) | 1,172<br>(91.14%) | 0.013 |
| 5 months n (%) | 118 (100.00%) | 24 (100.00%) | 34 (91.89%) | 88 (97.78%) | 979 (96.26%) | 1,243<br>(96.66%) | 0.08 |
| 6 months n (%) | 118 (100.00%) | 24 (100.00%) | 36 (97.30%) | 90 (100.00%) | 1,013<br>(99.61%) | 1,281<br>(99.61%) | 0.198 |

## Data Availability

All data produced in the present work are contained in the manuscript

## IVAC study group

Quentin Semanas, Antoine Oblette, Hadrien Regue, Geneviève Billaud, Martine Valette, Sophie Assant, Mary-Anne Trabaud, Bruno Pozzetto

